# PREDICTING LEVEL OF COMPLIANCE WITH INFECTION PREVENTION AND CONTROL PRACTICES AMONG HEALTHCARE WORKERS IN SOUTHERN NIGERIA USING ORDINAL LOGISTIC REGRESSION MODEL

**DOI:** 10.1101/2023.11.21.23298839

**Authors:** Maureen C Onyeloili

## Abstract

**Introduction:** Adherence to infection prevention and control guidelines is critical to improving the quality of hospital care based on their efficacy in reducing the occurrence of infections that compromise patients’ outcomes. However, the impact of predictors on IPC compliance among healthcare workers has not been adequately reported.

**Objectives:** This study aims to demonstrate the utility of the Ordinal Logistic regression model in identifying the impact of personal and organizational characteristics on health workers’ level of compliance with infection prevention and control at the University of Port Harcourt Teaching Hospital.

**Methods:** A cross-sectional study design using a self-administered questionnaire was adopted. A sample of 235 respondents was chosen using a proportionate stratified random sampling method. We analyzed data using descriptive statistics and the ordinal Logistic regression model.

**Result:** The study result shows that IPC compliance among Health workers in UPTH is high, 77%. Predictors of compliance were found to be age group 35-45years (AOR= 7.679, CI= 1.214 -48.577), training (AOR=0.401, CI: 0.189, 0.849), knowledge, (AOR= 0.45, CI: 0.207, 0.978), management support (AOR=0.45, CI 0.16, 0.968) as they were found to be statistically significant with the level of compliance with infection prevention and control.

**Conclusion:** There is relatively high Compliance with Infection Prevention and Control; this can be further improved through improved management commitment and increased surveillance of health workers.

**KEY MESSAGE:**

- **What is already known on this topic**: Several studies conducted have reported various factors affecting compliance with infection prevention and control among healthcare workers.
- **What this study adds:** The impact of the predictors on IPC compliance among healthcare workers has not been adequately reported. Hence, there is a methodology gap in the literature. The findings of this study gave insight and quantified the contribution of each predictor to Infection Prevention and Control compliance level, thus, deploying epidemiological and statistical methodology in Infection Prevention and Control studies.
- **How this study might affect research, practice or policy:** The study provides information that serves as a proactive guide on resource allocation and areas of improvement in the Infection Prevention and Control Compliance program for program evaluators, facility managers, health agencies, stakeholders, and other policymakers. It provides researchers with guidance on adopting epidemiological methodology in conducting evidence-based studies.

## Introduction

In the course of delivering care, healthcare workers are exposed to certain infections (1, 2). Patients seeking respite for various ailments are more vulnerable to these opportunistic infections (3). The infections are transmitted from patients through hands, equipment, supplies, and unhygienic practices. According to the Centre for Disease Control, Health-care Associated Originally, Healthcare Associated infections (HAIs) referred to illnesses contracted throughout the continuum of healthcare settings, not just those connected to a person’s time in an acute-care hospital (nosocomial). (e.g., Long-term care, home care, ambulatory care, end-stage renal disease facilities, amongst others)(4) These unanticipated infections develop during healthcare treatment and result in significant morbidity and mortality (5).Studies have shown that HAIs can culminate in prolonged hospital stays, a certain degree of disability, delayed response to treatment from increased resistance of microorganisms to antimicrobials, reduced productivity, increased costs for healthcare systems, negative cost implications for patients and their families, and avoidable death(6,7).

The burden of hospital-acquired infections is not known globally (8). However, the prevalence of healthcare-associated infections varies between 5.7% and 19.1 % in low and middle-income countries (9). There is a considerable burden of HAIs in developed countries. It reportedly affects 5% to 15% of hospitalized patients in regular wards and up to 50% or more of patients in intensive care units (ICUs) (10). This burden is felt many folds higher in low- and middle-income countries than in high-income countries (11). There is insufficient data to establish the prevalence of HAIs in Africa, and Nigeria, specifically, mainly because of the complex diagnosis of HAI and surveillance activities to guide interventions that require expertise and resources (12). Deficiencies in social and healthcare systems and economic problems account for an inadequate surveillance system. Overcrowding of patients and understaffing (6, 13) contribute to inadequate infection and control practices, the absence of infection control and prevention policies (14), absence of guidelines and trained professionals also aggravate the problem of inadequate prevention and control practices (15).

Effective infection prevention and control practices (IPC) have proven to reduce the burden of HAIs to the barest minimum (16, 17). Infection prevention and control are central to providing high-quality health care for patients and a safe working environment for those who work in healthcare settings (18). It is crucial to minimize the risk of the spread of infection to patients and staff in the hospital by complying with a good infection control program.

In Nigeria, there is a concerted effort by the National health body and healthcare facilities to develop infection control programs and strategies. The success of these programs depends to a large extent on compliance with these control guidelines. Adherence to infection prevention and control guidelines is critical to improving the quality of hospital care based on their efficacy in reducing infections that compromise healthcare delivery outcomes (19).

Healthcare workers (HCWs) play an important role in containing infectious diseases spread to patients and fellow health workers, who are also exposed to the infection in the facility. The emergence of several infectious diseases such as Sars-Cov-2 puts them at greater risk. It is imperative to remind health workers of their responsibility to abate infection vectors by complying with the laid down IPC guidelines. As reported in a study conducted by Lai et al., to effectively reduce the risk of particular infection transmission in healthcare institutions, HCWs must strictly comply with standardized preventive measures/guidelines and execute protective measures against droplet isolation, contact isolation, and air isolation aimed at breaking the chain of infection. (20)

IPC measures as recommended by WHO include hand hygiene, medical masks, use of personal protective equipment (PPE), single or cohort patients, sterilization of patient-care equipment, and linen, amongst others (21).

Healthcare workers in teaching and referral hospitals such as the University of Port Harcourt Teaching Hospital usually have the opportunity for training on infection prevention and control, which could increase their knowledge level. However, Compliance with IPC among HCWs was reportedly low (22). Healthcare providers have been found to demonstrate poor compliance with hand hygiene practices despite well-established guidelines for the prevention of healthcare-associated infections (23).

A study conducted on hand hygiene practice and awareness among health workers in selected facilities in Abuja, Nigeria, reported that 80% of the respondents agreed to hand washing before and after patient contact. Only 43.3% agreed to wash their hands after touching body fluids and taking off gloves because the hands did not come in direct contact with the patients. The study concluded that less than half of the respondents had excellent hand hygiene practices, while 37% had good practices, which was attributed to good knowledge but inadequate hand washing materials, inconveniently located sinks, and workload (24). In a study conducted by Haile et al. in Ethiopia, they reported that HCWs who had more frequent management support towards the safety environment in their institution were more likely to always comply with SPs than those who had less frequent support. Regarding organizational support in procurement of the Personal Protective Equipment (PPE), 5.4% PPE accessibility and 10% availability which contributed to low compliance with infection prevention and control was reported (25). The use of PPE consistently and effectively is necessary for protecting patients, healthcare workers, and their families. Personal protective equipment (PPE) can reduce the risk of infectious diseases by covering exposed body parts (26). Regarding needle stick injury, Hoffman discovered that the best way to protect against NSI is through safety devices. The study revealed that Needle-stick injury was reduced by 21.9%, the highest when safety devices were applied for blood withdrawal, peripheral venous catheters, and hypodermic needles (27).

Knowledge of Infection Control practices is becoming increasingly imperative with the emergence of infectious diseases, such as Coronavirus, Ebola, Lassa fever, avian influenza, severe acute respiratory syndrome, and the threat of bioterrorism (28).

The ecological models were adopted where the synergy of individual and organizational factors that play an essential role in infection control compliance was studied. The study looked at Infection Prevention and Control policies and guidelines, predictors of compliance, healthcare workers’ level of knowledge of the IPC, and the extent of training. It also tries to determine if compliance with IPC has any relationship with knowledge, training, socio-demographic factors of health workers, and how management support affects HCW’s Compliance with IPC.

Although several studies have investigated compliance on infection prevention and control. However, a literature search shows that only rare studies have been conducted on Ordinal regression models to determine the level of compliance to these infection control practices among healthcare workers. Hence, this study seeks to quantify the predicting factors that will enable Compliance with Infection Prevention and Control among healthcare workers in the University of Port Harcourt Teaching Hospital to ensure all other efforts do not end in a fiasco.

Therefore, this study is focused on predicting factors that enable Compliance with Infection Prevention and Control among healthcare workers using Ordinal Logistic Regression as a predictive tool. The study location is the University of Port Harcourt Teaching Hospital.

## MATERIALS AND METHODS

### Study Design

This research work aims to establish the Impact of the relationship between the health workers’ levels of compliance with infection prevention and control (IPC) practices and factors that affect the IPC compliance of these health workers. The study employed the institutional descriptive cross-sectional study design. An in-depth description of the IPC practices of healthcare workers at the study site was done using data obtained through a structured questionnaire. This study design is most appropriate to easily collect data at one time from study participants based on inclusion and exclusion criteria set for the study.

### Study Area

The study was carried out at the University of Port Harcourt Teaching Hospital (UPTH) located in Alakahia, Obio/Akpor Local Government Area of Rivers State, Nigeria. The hospital which is situated along the East-West Road of the city is a tertiary institution established in 1980. The institution is bounded on the east by Alakahia, on the West by Emohua, and the South by Aluu. Rivers State, Nigeria. It constantly draws patients from all the neighboring states of the oilLrich Niger Delta region; a catchment population that can be conservatively put at 10 million people.

### Study participants

The target population for this study was health workers employed in the University of Port Harcourt Teaching Hospital. The inclusion criteria were healthcare workers including doctors, nurses, pharmacists, laboratory scientist, and technologists units who had worked for at least one month at the institution and was willing to participate in the study. This population was targeted because they are in contact with the majority of patients attended to at the institution and their Infection Prevention and Control practices can either minimize or perpetuate the transmission of healthcare-associated infections (35).

All participants were aged 20 to 66 years. Their typical work day starts from 8 am to 4 pm after which the call duty starts till 8 am the following day. Participants were recruited between May 2021 to July 2021. However, HCWs on maternity leave, as well as those who have worked for less than 1 (one) month were excluded from the study.

### Study Variables

The outcome variable of the study was Healthcare workers’ level of Compliance with Infection Prevention and Control (IPC). The study’s independent variables identified from the literature were socio-demographic factors: (gender, marital status, age), individual factors (years of experience and knowledge of IPC), institutional factors (staff training on IPC and management commitment to IPC. The collection of data took place between June and July 2021.

### Sample Techniques/Sample Size Estimation

The proportionate stratified random sampling method was adopted. A simple random sampling technique was used to select respondents.

Sample Size Estimation

Using Cochran’s formula

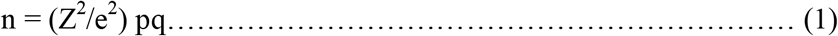

Where Z= 1.96 (at 95% confidence interval)

e= 5% (margin of error)

p= estimated proportion of attribute of interest (80%) (29)

n= 230.

A correction factor nf where the sample size is not up to 10,000 was adopted to compensate for respondents who could not return the study tool.

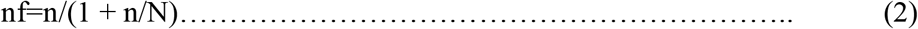

n= minimum sample size

n = 230

N = Total population of HCWs

N= 3542 (IPPIS Office UPTH)

nf = 219.

Nevertheless, the sample size was increased to 235 to compensate for non-returned, missing, unfilled, and inappropriately filled questionnaires.

### Data collection

Primary data were collected using a structured and self-administered questionnaire with the help of some medical house officers. The questionnaire was adapted from a previous study on “Compliance with Standard Precautions and Infection Prevention and Control among health workers (25) and further modified by the researchers to answer the research questions. Data collection was carried out from June 2021 to July 2021. Compliance with infection prevention and control was measured on a 3-point scale (1 = high compliance, 2 = moderate compliance, and 3 = poor compliance).

### Method of data analysis

Data entry was done with Excel, then cleaned and coded. Statistical analysis was carried out using IBM Statistical Package for Social Sciences (SPSS) version 20.0. Missing data were handled by substituting the missing data with the mean score of the variable (37). Descriptive statistics (percentages and frequencies) were calculated to demonstrate subjects’ demographics and characterize the distribution of variables. Ordinal logistic regression model analysis was carried out using SPSS to establish a relationship between the dependent and independent variables. Compliance, the dependent variable, was measured by 21 questions. We used a three-point response scale to measure compliance with the infection control and prevention guidelines at the workplace (Never = 0, Sometimes = 1, and Always = 2). Next, a summation of the 21 items was done. (Always=2, Seldom=1, Never= 0) total compliance=42 (2*21). Scores for each respondent were summed up and graded as high, moderate, or poor. High compliance: >= 28, moderate compliance= 14 – 27, poor compliance = <13. This scoring system has been used in an earlier study to investigate Compliance with IPC and Standard precautions among health workers in North-West Ethiopia (25). Knowledge, management support, and training as predictors were measured using the three-point response scale: Yes, No, Do not know. “Yes” signifies agreement, “No”, means not in agreement, and “do not know” implies uncertain or not sure.

Several independent variables were examined for possible association with compliance, including socio-demographics, knowledge, training and management support chosen a priori to reflect aspects of the underlying model guiding the study.

### Model Goodness of Fit

Model goodness of fit test was used to determine the possibility of predicted probabilities deviating from the observed, making it difficult for an ordinal logistic model to predict. Suppose the resulting p-value is less than the *p* =0.05 significant level. In that case, it is concluded that the predicted probabilities deviate from the observed in a way that the model could not predict. An unfit model could result from the incorrect link, omitted higher-order terms for variables in the model, or from the omitted predictor that is not in the model. If the deviation is statistically significant, then a new link function should be sorted, or the terms in the model will be changed (36).

For example, in this study, health workers’ specialty and ward of posting were automatically omitted from the model.

This study used the likelihood ratio test and the Pearson and Deviance Chi-Square statistic to measure the goodness of fit tests

The p-value of the Chi-square statistics from the table tells us if the variables added improved the model compared to the intercept-only (i.e., with no independent variables) model. The Pearson and Deviance Chi-Square statistics were used to determine if the model fits the data well.

Chi square statistics is given as;

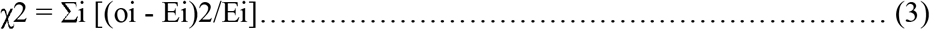

where ‘*oi*” represents the cases in category i’s observed value, and Ei represents the cases in category i’s expected value.

We computed Ordinal logistic regression analyses to identify variables having a significant association with the dependent variable. An odds ratio with a 95% confidence interval determines the strength of the association between dependent and independent variables. Having a *p-value* of less than 0.05 in the ordinal logistic regression model was considered a significantly associated variable.

### The ordinal regression model equation is given by

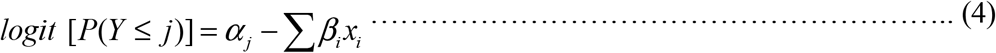

j = 1, 2… p (p is the number of levels of the dependent variable minus 1)

i = 1, 2 … m (m is the number of the predictor variables)

(α; constant; β; coefficient; χ; explanatory variable)

A link function that maps probabilities to the real line is denoted by (*g)*. The cumulative probabilities are converted into a linear function of the predictor variables using the link function. The cumulative response probabilities’ *(Yij)* logit transformation is provided by;

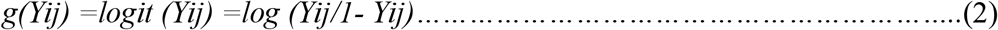

The explanatory variables either increase or decrease the likelihood of a response in category j.

### Ethical Concerns

Ethical clearance was obtained from the ethical review committee of the University of Port Harcourt Ethical Review Committee (certificate number. UPH/CEREM/REC/MM75/090) and the University of Port Harcourt Teaching Hospital Research Ethics Committee (certificate number, UPTH/ADM/90/S.II/VOL.XI/1244). Written informed consent was obtained from each study participant after explaining how they would take part in the study and if any involvement was required after they completed the consent. Anyone not willing to participate in the study was given the full right not to participate. The researcher used codes rather than names and kept the questionnaires sealed in an envelope to ensure confidentiality

## Result

This study had a total of 332 respondents (health workers) who were administered the questionnaires. According to the response return rate based on the questionnaires that were returned and duly filled, 245 respondents participated in the study with 10 questionnaires incompletely filled, implying that the response return rate of 70% was achieved. The effective response rates were determined by the ratio of survey questionnaires completed to the total number of survey questionnaires issued at the study location multiplied by 100. This response return rate was achieved because of the researcher’s constant follow-up on the study participants. The socio-demographic characteristics of the participants was shown in Table 1 (Appendix).

Figure 1 is a pie chart showing high IPC compliance, but the study is concerned with the 23% that reported not being highly IPC compliant.

**Figure 1.**
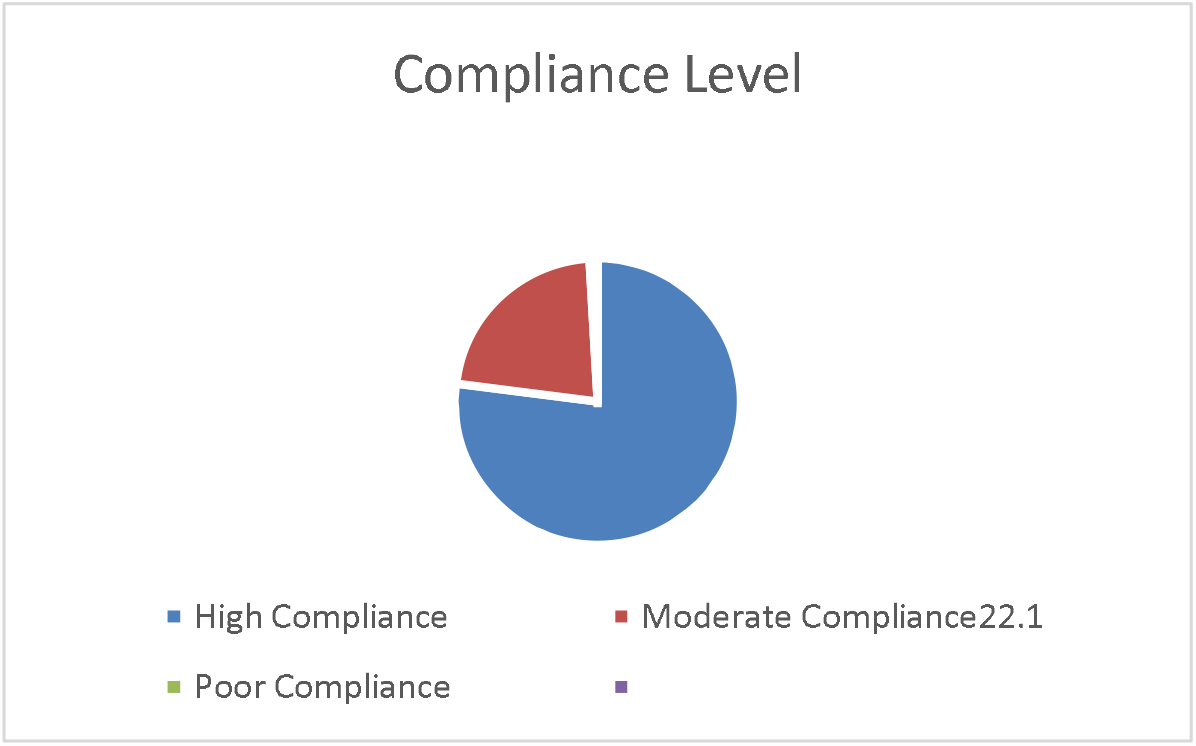
A pie chart showing the level of Compliance with IPC among Health workers in UPTH.

Table 2 shows the Parallel Regression Assumption test using the Test of Parallel Lines in SPSS.

**Table 2.** Test of parallel lines.

| Model | -2 Log Likelihood | Chi-Square | Df | Sig. |
| --- | --- | --- | --- | --- |
| Null Hypothesis | 139.545 |  |  |  |
| General | 130.079 | 9.466 | 10 | .489 |

The Test of parallel lines in table 2 above is not statistically significant, (p=.489), hence, we conclude that the proportionality odds assumption, also known as the parallel regression assumption, is satisfied. This implies that we can proceed with the ordinal logistics regression analysis

The regression coefficient of the variables are shown in Table 3 (Appendix). For our parameter estimation, recall from equation (4), the Logit model is given as: *logit*[*P*(*Y* < *j*)]=*α*_*j*_ −∑*β*_*i*_*x*_*I*_ Therefore, the equation of the model is given as:

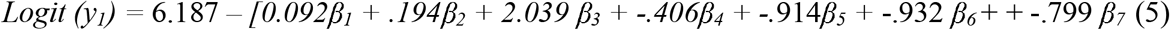

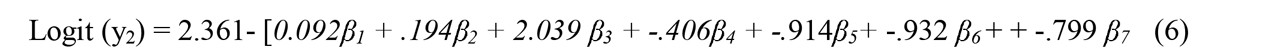

**Table 3:**
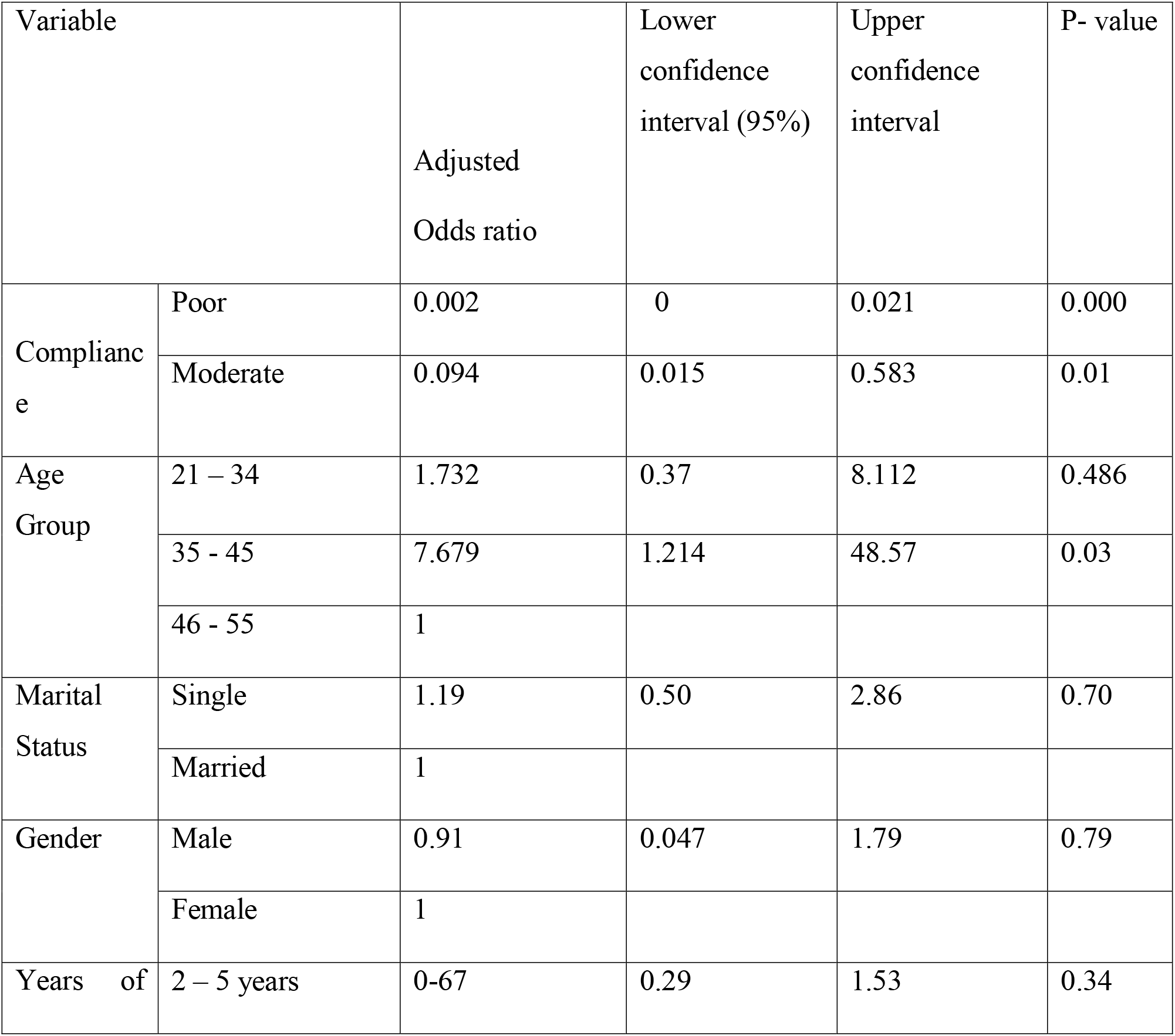
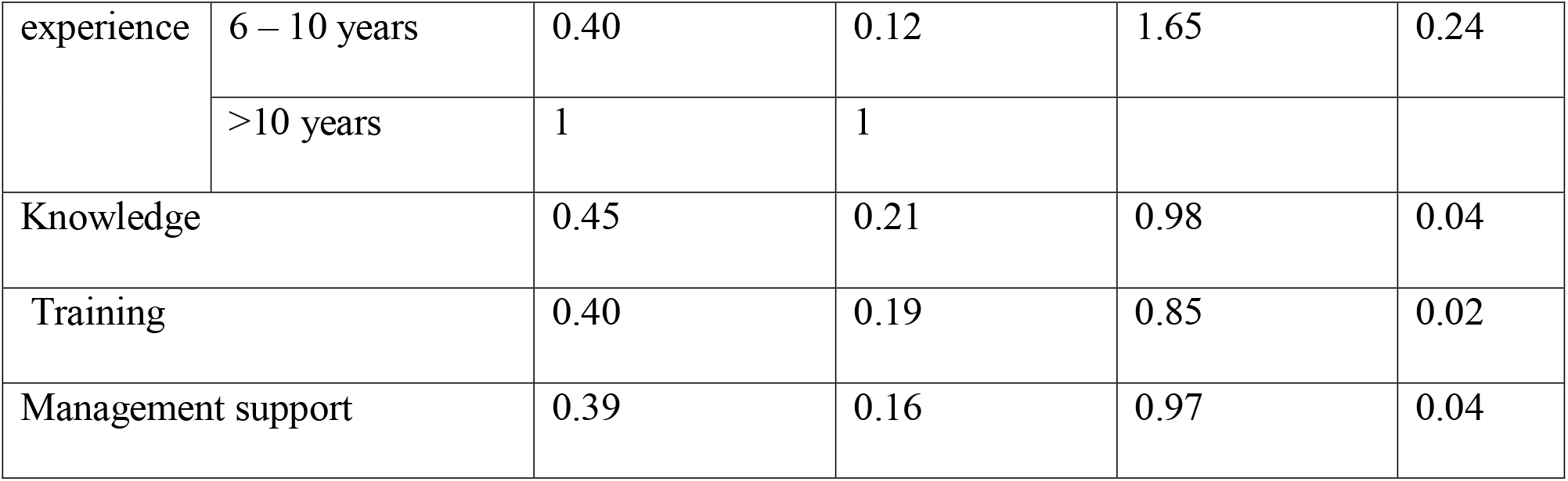
Odds ratio and confidence Intervals of the Variables.

As seen from Table 3 above only age group 35-45, knowledge, training, and management support were found to be statistically significant (*p=*.*05*), hence are the only predictors of IPC compliance in this study. The table shows that the confidence intervals for gender and years of experience include one which implies that they are not significant predictors of IPC compliance among health workers.

## DISCUSSIONS

This research work aims to utilize ordinal logistic regression to illustrate the relationship between the level of infection Prevention and Control IPC compliance and its predictors among healthcare workers at the University of Port Harcourt. A sample of 235 students was obtained using the stratified random sampling method. The level of compliance with infection prevention and control among health workers is an ordinal response categorized according to the level of Compliance with IPC (High Compliance, Moderate Compliance, Low Compliance). Included in the model are five predictors of IPC compliance, namely; Socio-demographic characteristics: gender, age, Years of experience, IPC Training, Knowledge of IPC, and management commitment to IPC. The model goodness of fit test was carried out, and the result shows that the model was a good fit. The results of the ordinal regression in Table 3 show that all considered variables except socio-demographic factors were found to be significant predictors of Compliance with IPC, evidenced in the p-value being more significant than 0.05 (p>0.05).

The results in Table 4 also show that when the IPC knowledge increases by a unit the odds of the IPC compliance level of the health worker being in a poor or moderate-level versus high level decreases by 45%, given that any other variable is held constant Most of the health workers that participated in this study are between 21 – 34 years; they still have long years to add to work. There is a great need to protect young workers from hospital-acquired infections. Previous studies reported a similar age group as the most dominant age bracket among the study participants (30,31),

In line with this study, American and Onwube documented no statistical significance between gender, marital status, years of work of health workers, and IPC compliance in a similar study conducted in Northern Nigeria (31).

The study findings show that the level of Compliance with IPC among UPTH health workers is high (77%). Our findings are congruent with the report in a Ghanaian study where the compliance is high, calculated from high hand hygiene compliance and High PPE compliance (95.5 % respectively) (32). However, contrary to our findings, a study conducted in China reported sub-optimal compliance with infection prevention and control relating to patient handling (54%) and invasive procedures (46%) (20) and reported a low level of Compliance to IPC among HCWs. The contrast can be linked to Healthcare workers in China being unaware of the pandemic, which started in China. China had not embraced infection prevention and control to contain the menace of the pandemic as the study was conducted in the middle of the pandemic. Also, the study findings are contrary to a similar study conducted in Southern Nigeria by Uvieroghene and Best, where they reported deficient practice of IPC despite reporting high knowledge (24). This is an indication that knowledge most times does not translate to practice. The difference can be attributed to the concerted effort made by the Ministry of Health, the Nigeria Centre for Disease Control, and other health agencies to increase awareness and improve infection prevention and control following the upsurge of the COVID-19 pandemic in 2020. Another similar study conducted in the facility reported low compliance with hand washing (24)

The study further revealed that half of the health workers reported being trained on IPC. The study is congruent with the study conducted by Keita et al. in Guinea. A high IPC score was positively associated with the number of trained staff; IPC-trained workers were eight times as likely to have a high IPC compared to facilities with no trained workers (33). A similar result was found in a previous study conducted in Southern Ethiopia, where training has increased the odds of Compliance with IPC and standard precautions, and the health workers reported an adjusted odds ratio of 3.99(1.46, 10.9) for Infection Prevention training(25).

On knowledge, the study findings agree with Ogoina et al. in a study in Nigeria that documented above 90% of health workers’ knowledge of IPC. However, the median practice score was 50.8% showing that knowledge did not translate to practice. However, most HCWs had poor knowledge of injection safety and complained of inadequate resources to practice standard precautions (34). Another previous study reported high knowledge of IPC among health workers (30). A contrasting report was found in a previous study in Northern Nigeria (31). The difference can be attributed to the different years of the study and literacy margin in Nigeria Northern Nigeria.

One significant finding is the management support which is reported to be 34.9%. Poor management is a disturbing finding for a hospital like the University of Port Harcourt Teaching Hospital, which has an Infection Prevention and Control Unit who are supposed to know the causal relationship between management commitment and IPC compliance. The facility should display leadership by providing needed support regarding availability and access to PPE, Hand hygiene equipment, and adequate training to health workers. Our findings are in line with a similar study in Ethiopia. HCWs who had adequate management support towards a safe working environment at the institution were 2.23 times more likely to be compliant than those who had less frequent management support. Also, the finding is consistent with a study in Northern Nigeria where non-compliance to IPC is attributed to poor management support. According to the study, a majority (98.6%) of the respondents reported that the primary reason for non-compliance to universal precautions is the non-availability of IPC equipment (31).

### Limitations of study

The short period of the researcher’s program did not allow for proper in-house monitoring of health workers carrying out these IPC practices but was based on self-reported compliance. Hence, there is a tendency for reporting bias which was taken care of by detailed and straightforward questions in the questionnaire. Real-time behavioral compliance may not have been captured. The researcher only based her findings on responses elicited from the questionnaire. The research tool was developed by the authors and maybe subject to bias. But the authors have pre-tested and validated on a smaller sample. Further investigation should consider the real-time practice of health workers’ compliance to IPC.

## Conclusion

The ordinal logistic regression model analysis results show the ordered category of Health workers’ IPC compliance level at the University of Port Harcourt Teaching Hospital. We can conclude that knowledge, management support, training, and a specific age category are the influential factors that affect the IPC Compliance level.

The reported compliance level was high, which was attributed to a combination of predictors. Inadequate management support was the main reason for the poor compliance reported.

## Data Availability

All data produced in the present study are available upon reasonable request to the authors

